# PCSK9 inhibition in myeloid cells enhances cardioprotection beyond its LDL cholesterol-lowering effects

**DOI:** 10.1101/2024.08.27.24312680

**Authors:** Shin Hye Moon, Hyo Won Ki, Na Hyeon Yoon, Katherine I. Chung, Huiju Jo, Jing Jin, Sejin Jeon, Seong-Keun Sonn, Seungwoon Seo, Joowon Suh, Hyae Yon Kweon, Yun Seo Noh, Won Kee Yoon, Seung-Jun Lee, Chan Joo Lee, Nabil G. Seidah, Sung Ho Park, Goo Taeg Oh

**Affiliations:** Heart-Immune-Brain Network Research Center, Department of Life Sciences, Ewha Womans University, Seoul, 03760, Republic of Korea; Department of Biological Sciences, Ulsan National Institute of Science & Technology (UNIST), Ulsan, 44919, South Korea; Department of Vaccine Biotechnology, Andong National University, Andong, Republic of Korea; Imvastech Inc., 52 Ewhayeodae-gil, Seodaemun-gu, Seoul, 03760, Republic of Korea; Korea Research Institute of Bioscience & Biotechnology, Laboratory Animal Resource Center, 30 Yeongudanji-ro, Ochang-eup, Cheongwon-gu, Cheongju-si, Chungcheongbuk-do, 28116, Korea; Division of Cardiology, Severance Cardiovascular Hospital, Yonsei University College of Medicine, 50-1, Seoul, 03722, Republic of Korea; Laboratory of Biochemical Neuroendocrinology, Montreal Clinical Research Institute (IRCM), Montreal, Quebec H2W 1R7, Canada

**Keywords:** coronary artery disease, lipid homeostasis, PCSK9 inhibitors, cardiac macrophages, activator protein-1, vascular endothelial growth factor C

## Abstract

**BACKGROUND:** Circulating levels of proprotein convertase subtilisin/kexin type 9 (PCSK9), which regulates plasma cholesterol content by degrading LDL receptor, are correlated with the risk of acute myocardial infarction (AMI). Recent studies suggested that PCSK9 improves cardiac function beyond its effects on LDL cholesterol levels after cardiac ischemic injury, but its precise mechanism remains unclear.

**METHODS:** We examined the interrelationship and functional significance of PCSK9 and cardiac myeloid cells in ischemic hearts from AMI-induced *Pcsk9^-/-^* and *Lyz2^cre^Pcsk9^fl/fl^*mice, as well as in serum samples from coronary artery disease (CAD) patients treated with PCSK9 antibodies (Ab). Single-cell RNA sequencing (scRNA-seq) was conducted to identify heterogenous cardiac macrophage clusters and to investigate the impact of adaptive remodeling due to PCSK9 deficiency during AMI. Additionally, the regulatory effect of the myeloid-PCSK9/VEGF-C pathway was assessed *in vitro* as a potential therapeutic strategy.

**RESULTS:** Our study demonstrated that PCSK9 deficiency induces diverse changes in myeloid cells and macrophages, potentially offering cardiac protection following AMI, irrespective of LDL cholesterol homeostasis. The scRNA-seq identified a subset of PCSK9-dependent cardiac macrophages (PDCMs) enriched in activator protein-1 (AP-1)–related pathways, functioning as reparative macrophages. These PDCMs were shown to enhance vascular endothelial growth factor C (VEGF-C) secretion and activate Akt signaling in cardiac endothelial cells, leading to improved cardiac remodeling. Notably, CAD patients treated with PCSK9 inhibitors exhibited increased numbers of myeloid cells with PDCM-like features, including elevated VEGF-C levels, consistent with our findings in mice.

**COUNCLUSIONS:** Targeting PCSK9 in myeloid cells could offer cardioprotective effects by increasing AP-1 activity and VEGF-C expression of PDCMs, presenting a novel approach to preventing cardiac dysfunction in AMI. This strategy could expand the clinical use of existing PCSK9 inhibitors beyond just lowering LDL cholesterol.

**Clinical Perspective:** *What is New?:*

- Myeloid-PCSK9 deficiency attenuated cardiac dysfunction post-acute myocardial infarction (AMI) without affecting plasma lipid levels. These findings position PCSK9 as a novel immune regulator of macrophages, revealing functions independent of its role in LDL cholesterol regulation.
- We demonstrated PCSK9-dependent cardiac macrophages (PDCMs) that play a reparative role under ischemic conditions influenced by PCSK9, using single-cell RNA sequencing (scRNA-seq) of CD45^+^ leukocytes following AMI.
- Strong enrichment of AP-1 family proteins in PDCMs led to reparative VEGF-C signaling in endothelial cells and improved cardiac remodeling, independent of PCSK9’s conventional role in cholesterol homeostasis.
- In coronary artery disease (CAD) patients, PCSK9 inhibition augmented myeloid cell populations towards a reparative phenotype and elevated VEGF-C levels, aligning with our findings in mice.

*What Are the Clinical Implications?:*

- Myeloid-derived PCSK9 is pathobiologically significant, directly influencing immune functions and contributing to cardiac remodeling after AMI, suggesting that targeting myeloid-specific PCSK9 could be a valuable therapeutic approach.
- Given that the reparative effects of PCSK9 inhibitors on macrophages are preserved in CAD patients, this strategy could broaden the clinical applications of existing PCSK9 inhibitors beyond LDL cholesterol regulation.

## Introduction

Acute myocardial infarction (AMI) is a leading global cause of mortality, and it is characterized by a pronounced inflammatory response in the heart and blood. This response exacerbates cell death, impairs heart function, and promotes ventricular adverse remodeling.^1^ Despite available therapeutic targets, AMI carries high risks of heart failure^2^ and mortality, emphasizing the need for further research into immune cell mechanisms to improve treatment strategies.

Proprotein convertase subtilisin/kexin type 9 (PCSK9) is primarily known to increase circulating LDL cholesterol (LDL-c) levels by promoting LDL receptor (LDLR) degradation.^3–5^ However, previous studies revealed that PCSK9 inhibition and deletion significantly reduce cardiac dysfunction^6^, suggesting a potential role of immune cells in the reparative phase within the ischemic heart.^7^ Macrophages are highly active cell types throughout all stages after AMI, including the inflammatory, tissue repair, and cardioprotective phases.^8,9^ Therefore, understanding the influence of PCSK9 on macrophages is essential for diagnosing and treating cardiovascular diseases, including myocardial infarction and atherosclerosis.

In this study, we revealed that PCSK9 deletion induces macrophage heterogeneity with reparative characteristics against AMI, defining them as PCSK9-dependent cardiac macrophages (PDCMs). We demonstrated that PDCMs are crucial in the reparative phase following myocardial infarction (MI), promoting cardiac healing and reducing adverse remodeling. Mechanistically, activated activator protein-1 (AP-1) in PDCMs induces the production of vascular endothelial cell growth factor C (VEGF-C) to activate Akt signaling in endothelial cells and facilitate cardiac repair and regeneration. Furthermore, we observed that PCSK9 antibody-treated patients with coronary artery disease (CAD) have PDCM-like features in blood myeloid cells. Collectively, we verified that targeting PCSK9 in myeloid cells (myeloid-PCSK9) enhances reparative tissue remodeling through the AP-1/VEGF-C/p-Akt axis, promoting protective effects from PDCMs to endothelial cells in ischemic hearts, irrespective of LDL-c levels.

## Methods

Data, analytical methods, and study materials that support the findings of this study are available from the corresponding author on reasonable request. Detailed methods are described in the Data Supplement.

### Mice Study Approval

All animal experiments were approved by the Institution Animal Care and Use Committees (IACUC) of Ewha Womans University, Seoul, Korea (IACUC No. 19-004, and 21-051).

### Human Samples

The Institutional Review Board of Severance Hospital (Seoul, Korea; IRB: 4-2023-0509) approved the protocol for collecting human blood samples. Written informed consent was provided by all participants before enrollment. Identifying information was removed from all samples before analysis for strict privacy protection. All experiments conducted with human blood samples were performed in accordance with the relevant guidelines and regulations.

### Data availability

scRNA-seq data for this project have been deposited at NCBI’s Gene Expression Omnibus (GEO) and GSE number is pending.

### Statistical Analysis

All statistical analyses were conducted using GraphPad Prism 8 (GraphPad). Comparisons between two groups were performed using a paired or unpaired two-tailed Student’s *t*-test or nonparametric two-tailed Mann–Whitney U-test, whereas comparisons among three or more groups were performed using one-way ANOVA. Data were normalized to the control group to reduce batch effects in some experiments. Mouse survival was measured by the number of deaths after LAD ligation, and the data was analyzed using the Kaplan–Meier method. The results were presented as the means ± SEM, and *P* < 0.05 was considered statistically significant.

## Results

### Myeloid-PCSK9 deficiency inhibits myocardial damage without affecting lipid homeostasis

Previous studies suggested that PCSK9 exacerbates cardiac dysfunction post-AMI^6,10^, with high circulating PCSK9 levels associated with an increased CAD risk.^11–14^ Although PCSK9 is primarily secreted by hepatocytes, it is also expressed by cardiac macrophages, endothelial cells, and smooth muscle cells.^15^ Thus, PCSK9 inhibition has been demonstrated to reduce CAD risk by affecting immune cell populations. Several reports found that PCSK9 blockade attenuates cardiovascular diseases such as atherosclerosis^16,17^ and aortic aneurysm.^18^ However, there is a lack of cell-specific studies on PCSK9 in myocardial infarction, particularly regarding the relationship between immune cells and PCSK9. To elucidate the role of PCSK9 in immune cells during heart injury, we subjected conventional PCSK9-deficient (*Pcsk9^−/−^*) and myeloid cell-specific PCSK9 knockout (*Lyz2^cre^Pcsk9^fl/fl^*) mice to left anterior descending (LAD) coronary artery ligation or sham surgery. Cardiac function and immune cell populations were assessed by echocardiography and flow cytometry, respectively, on days 3 and 7 post-AMI (Figure 1A and Figure IA in the Data Supplement). As expected, PCSK9 deficiency reduced plasma cholesterol levels and significantly improved mortality rates, although there was no significant change in the heart weight to body weight (HW/BW) ratio (Table I and Figure IB-ID in the Data Supplement). After MI, PCSK9 deficiency also reduced infarct size, attenuated wall thinning, and improved cardiac contractile function, indicating a protective effect (Figure IE, IF and Table II in the Data Supplement). These findings were associated with decreased numbers of total leukocytes and macrophages in the PCSK9-deficient infarcted heart, but no differences in the counts of other immune cells were recorded (Figure IG and IH in the Data Supplement).

**Figure 1.**
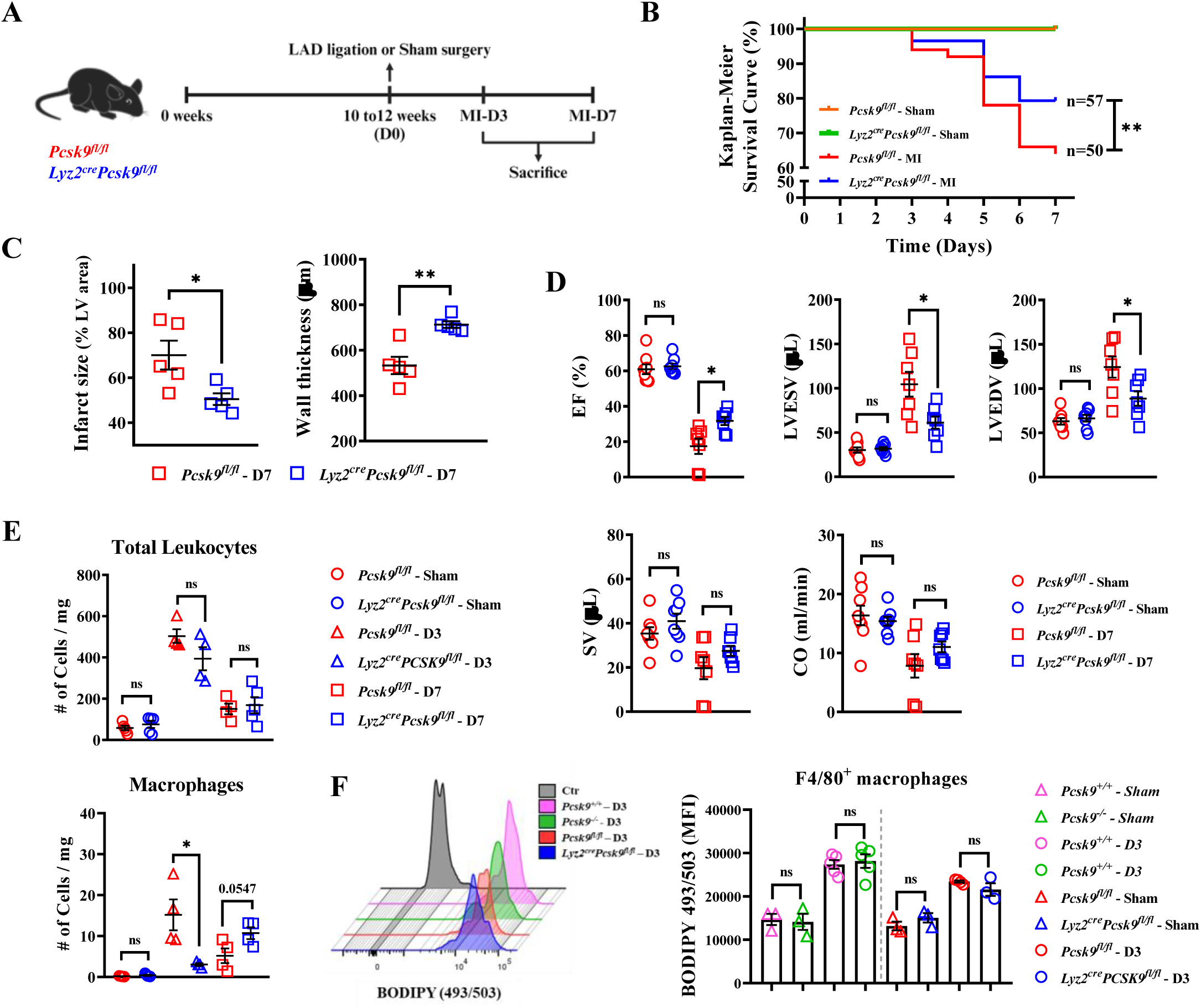
PCSK9 exacerbates myocardial injury by modulating cardiac macrophages in the ischemic heart. **A,** Experimental design: 10–12-week-old male *Pcsk9^fl/fl^* and *Lyz2^cre^Pcsk9^fl/fl^* mice were subjected to LAD coronary artery ligation to induce AMI. Cardiac function was assessed using echocardiography and immune cell analysis by flow cytometry on day 3 or 7 post-surgery and then sacrificed. **B,** Mortality rates of male *Pcsk9^fl/fl^* and *Lyz2^cre^Pcsk9^fl/fl^* mice up to 7 days post-surgery. **C,** Percent infarct size to the LV area (**Left**; *n* = 5). LV wall thickness (µm) post-LAD ligation in the MI-D7 groups of male *Pcsk9^fl/fl^* and *Lyz2^cre^Pcsk9^fl/fl^* mice (**Right**; *n* = 5). **D,** Echocardiographic assessment of ejection fraction (EF), LV end-systolic volume (LVESV), LV end-diastolic volume (LVEDV), stroke volume, and cardiac output. **E,** The counts of the indicated myeloid cells in the sham (*n* = 5), MI-D3 (*n* = 4), and MI-D7 (*n* ≥ 4) groups of *Pcsk9^fl/fl^* and *Lyz2^cre^Pcsk9^fl/fl^* mice. **F,** Flow cytometry revealing BODIPY (493/503) staining of F4/80 macrophages from the sham (*n* = 3) or MI-D3 (*n* = 5) hearts of *Pcsk9^+/+^* and *Pcsk9^−/−^* mice, and from sham (*n* =3) or MI-D3 (*n* ≥ 3) hearts of *Pcsk9^fl/fl^* and *Lyz2^cre^Pcsk9^fl/fl^* mice. For **C–F,** each dot represents one mouse. All data are presented as the mean ± SEM. ns, P > 0.05; \**P* < 0.05, \*\**P* < 0.01.

Assessment of the response of macrophages to AMI indicated that PCSK9 plays a pivotal role in macrophages during cardiac ischemia. Substantiating this finding, we confirmed that *Pcsk9* mRNA levels were considerably reduced in CD11b^+^ myeloid cells isolated from the sham-operated or post-AMI hearts of *Lyz2^cre^Pcsk9^fl/fl^*mice (Figure IIA in the Data Supplement). Interestingly, myeloid cell-specific PCSK9 deficiency improved the survival rate and reduced the incidence of cardiac rupture post-AMI without affecting the HW/BW ratio (Figure 1B, Figure IIB and IIC in the Data Supplement). Masson’s trichrome staining and echocardiography illustrated that myeloid-PCSK9 deletion reduced infarct size, maintained left ventricular (LV) wall thickness, and improved cardiac function, as evidenced by enhanced EF and reduced LV end-systolic volume, LV end-diastolic volume, and LV end-diastolic diameter (Figure 1C and 1D, Figure IID and IIE, and Table III in the Data Supplement). Additionally, flow cytometry revealed that myeloid-PCSK9 deficiency reduced cardiac macrophage counts, whereas the counts of other immune cells were not changed after AMI (Figure 1E and Figure IIF in the Data Supplement). These results revealed that targeting myeloid-PCSK9 attenuates cardiac injury and improves survival following AMI.

PCSK9 influences plasma cholesterol homeostasis by increasing LDL-c levels through clearing LDLR.^19,20^ Additionally, macrophages play a significant role in reverse cholesterol transport and the development of cardiovascular diseases.^21,22^ To investigate the association between PCSK9 and macrophages in plasma cholesterol homeostasis, we evaluated lipoprotein uptake and plasma lipid levels. Flow cytometry using the fluorescence probe BODIPY (493/503) demonstrated that neither sham surgery or cardiac ischemia affected the lipid droplet accumulation of F4/80^+^ macrophages in *Pcsk9^−/−^* and *Lyz2^cre^Pcsk9^fl/fl^*mice compared with their controls (*Pcsk9^+/+^* and *Pcsk9^fl/fl^*, respectively, Figure 1F). Plasma lipid profiles, including total cholesterol (CHO), HDL cholesterol (HDL-c), and LDL-c levels, were not altered in mice with myeloid-specific PCSK9 deficiency (Table 1). These data imply that myeloid-PCSK9 deficiency does not impact plasma cholesterol homeostasis. Overall, our findings indicate that myeloid-PCSK9 exacerbates myocardial injury in the ischemic state independent of its effects on LDL-c levels.

**Table 1.**
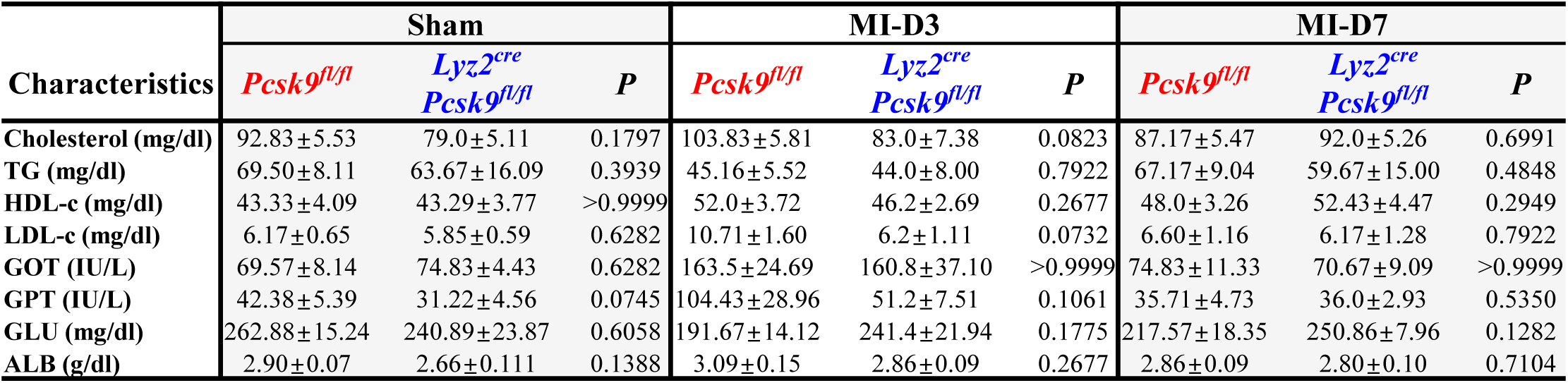
Myeloid-specific PCSK9 deficiency does not affect plasma lipid levels after LAD ligation. Plasma lipid profiles were assessed in the sham (*n* = 6), MI-D3 (*n* ≥ 5), and MI-D7 (*n* = 6) groups of *Pcsk9^fl/fl^* and *Lyz2^cre^Pcsk9^fl/fl^* mice. All data are presented as the mean ± SEM. An unpaired two-tailed Student’s *t*-test was used for statistical analysis. *P*-values are indicated for comparisons between *Pcsk9^fl/fl^ vs. Lyz2^cre^Pcsk9^fl/fl^* mice within each group. TG, Triglyceride; HDL-c, high-density lipoprotein cholesterol; LDL-c, low-density lipoprotein cholesterol; GOT, glutamic oxaloacetic transaminase; GPT, glutamic pyruvic transaminase; GLU, glucose; ALB, albumin. ns, *P* > 0.05

| Characteristics | Sham |  |  | MI-D3 |  |  | MI-D7 |  |  |
| --- | --- | --- | --- | --- | --- | --- | --- | --- | --- |
|  | *Pcsk9<sup>fl/fl</sup>* | *Lyz2<sup>cre</sup>*<br>*Pcsk9<sup>fl/fl</sup>* | *P* | *Pcsk9<sup>fl/fl</sup>* | *Lyz2<sup>cre</sup>*<br>*Pcsk9<sup>fl/fl</sup>* | *P* | *Pcsk9<sup>fl/fl</sup>* | *Lyz2<sup>cre</sup>*<br>*Pcsk9<sup>fl/fl</sup>* | *P* |
| Cholesterol (mg/dl) | 92.83±5.53 | 79.0±5.11 | 0.1797 | 103.83±5.81 | 83.0±7.38 | 0.0823 | 87.17±5.47 | 92.0±5.26 | 0.6991 |
| TG (mg/dl) | 69.50±8.11 | 63.67±16.09 | 0.3939 | 45.16±5.52 | 44.0±8.00 | 0.7922 | 67.17±9.04 | 59.67±15.00 | 0.4848 |
| HDL-c (mg/dl) | 43.33±4.09 | 43.29±3.77 | >0.9999 | 52.0±3.72 | 46.2±2.69 | 0.2677 | 48.0±3.26 | 52.43±4.47 | 0.2949 |
| LDL-c (mg/dl) | 6.17±0.65 | 5.85±0.59 | 0.6282 | 10.71±1.60 | 6.2±1.11 | 0.0732 | 6.60±1.16 | 6.17±1.28 | 0.7922 |
| GOT (IU/L) | 69.57±8.14 | 74.83±4.43 | 0.6282 | 163.5±24.69 | 160.8±37.10 | >0.9999 | 74.83±11.33 | 70.67±9.09 | >0.9999 |
| GPT (IU/L) | 42.38±5.39 | 31.22±4.56 | 0.0745 | 104.43±28.96 | 51.2±7.51 | 0.1061 | 35.71±4.73 | 36.0±2.93 | 0.5350 |
| GLU (mg/dl) | 262.88±15.24 | 240.89±23.87 | 0.6058 | 191.67±14.12 | 241.4±21.94 | 0.1775 | 217.57±18.35 | 250.86±7.96 | 0.1282 |
| ALB (g/dl) | 2.90±0.07 | 2.66±0.111 | 0.1388 | 3.09±0.15 | 2.86±0.09 | 0.2677 | 2.86±0.09 | 2.80±0.10 | 0.7104 |

### PCSK9 alters cardiac macrophage heterogeneity after AMI

Single-cell RNA sequencing (scRNA-seq) was conducted to identify changes in macrophage heterogeneity and understand the effects of PCSK9 deficiency on cardiac repair. Analysis of sorted CD45^+^ cells from noninfarcted and infarcted hearts revealed eight transcriptionally distinct immune cell populations by representative markers (Figure IIIA-IIIC in the Data Supplement). Ischemic damage led to significant increases in monocyte/macrophage (Mo/Mac) and neutrophil counts, whereas those of B cells, T cells, and NK cells were decreased (Figure IIID and IIIE in the Data Supplement). These data indicate that cardiac ischemia dramatically alters immune cell heterogeneity, with a significant involvement of macrophages, as expected.

To further characterize the role of macrophage-specific PCSK9 deficiency in cardiac regeneration, we defined subpopulations of Mo/Mac and Mo/Mac/DC clusters through clustering analysis. Eleven clusters were identified, including five macrophage clusters, three monocyte clusters, and two DC clusters (Figure 2A and Figure IVA in the Data Supplement). Notably, two macrophage clusters (Lyve1^+^ and CX_3_CR1^+^MHCII^+^) were significantly restored in *Pcsk9^−/−^* hearts compared with the findings in *Pcsk9^+/+^* hearts on MI-D3 (Figure 2B, 2C and Figure IVB in the Data Supplement). Both clusters, which are known for their reparative roles, including angiogenic and wound healing characteristics, were previously identified in published studies.^23–25^ To confirm the presence of reparative macrophages in the setting of PCSK9 deficiency, bone marrow-derived macrophages (BMDMs) from *Pcsk9^−/−^* mice were treated with IL-4 and IL-13 to generate PCSK9-deficient BMDMs (BMDM(IL-4/IL-13)^ΔPCSK9^). The absence of PCSK9 increased the mRNA levels of anti-inflammatory genes (*Arg1*, *Mrc1*, *Tgf-β*) in BMDM(IL-4/IL-13)^ΔPCSK9^ but not BMDM(IL-4/IL-13)^Ctr^, whereas *Il-10* expression was not significantly affected (Figure IVC in the Data Supplement). These clusters, which we termed PDCMs, are believed to facilitate tissue remodeling and repair in AMI. Consistent with the results of scRNA-seq analysis, elevated expression of Lyve1 and CX_3_CR1 in F4/80^+^ macrophages (PDCMs) was observed in the ischemic area of myeloid-specific PCSK9-deficient mice (Figure 2D-2G). Flow cytometry revealed increases in PDCM counts in the hearts of *Lyz2^cre^Pcsk9^fl/fl^* mice compared with the findings in *Pcsk9^fl/fl^* mice on MI-D7 (Figure IVD in the Data Supplement). Together, these results suggest that PCSK9 alters macrophage heterogeneity in ischemia, and activated PDCMs, in the absence of PCSK9, may promote reparative processes at the site of cardiac ischemia.

**Figure 2.**
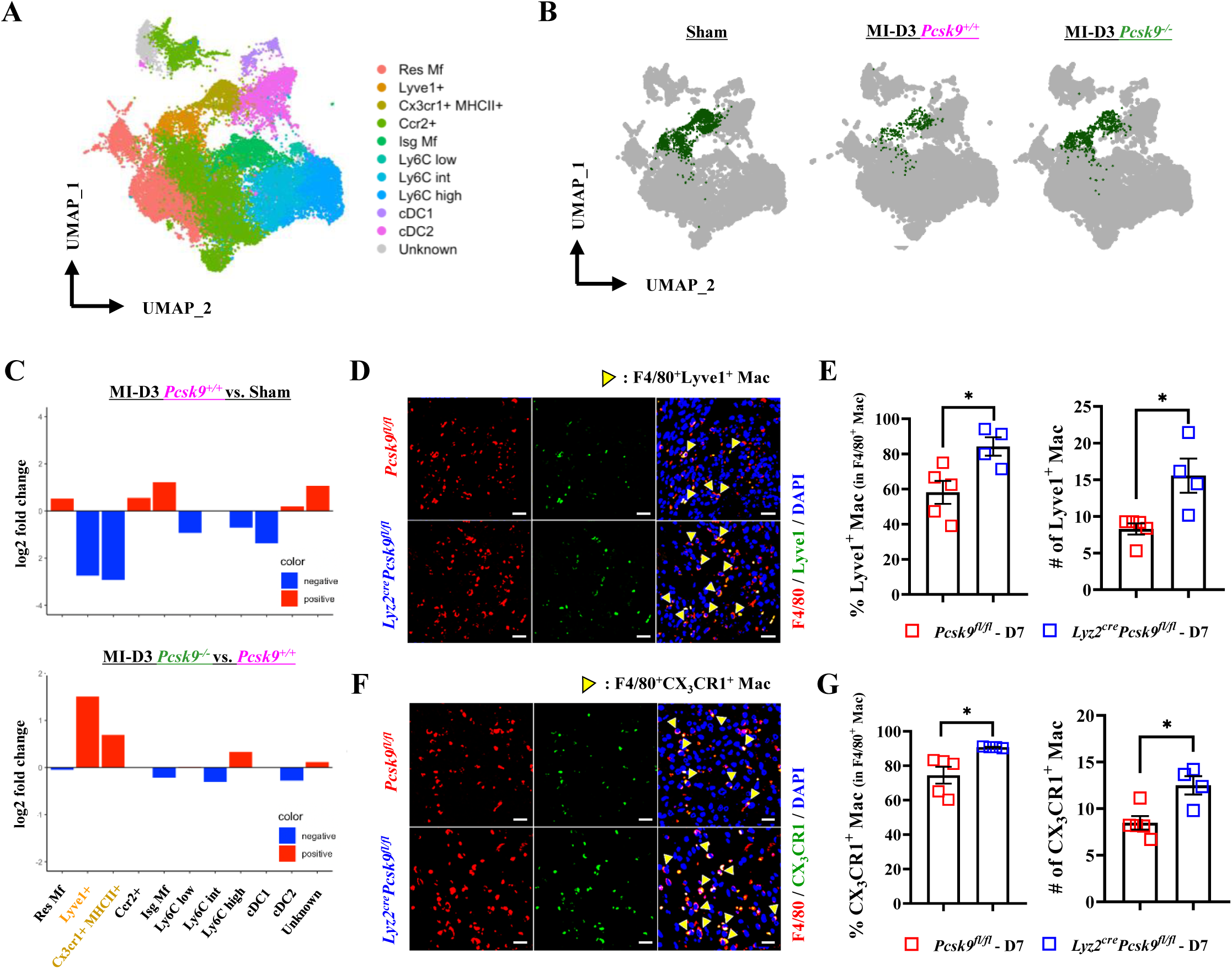
scRNA-seq analysis reveals PCSK9-dependent Cardiac Macrophages (PDCMs). **A,** Among the eight clusters (Figure IIIB), only macrophage-related clusters (Mo/Mac and Mo/Mac/DC) were subclustered, resulting in 11 newly defined subclusters. UMAP plots present these 11 subclusters and annotations of cells identified in ischemic hearts from the sham, MI-D3 *Pcsk9^+/+^*, and MI-D3 *Pcsk9^−/−^* groups. **B,** UMAP plots specifically highlighting PDCM clusters (Lyve1+ and Cx3cr1+MHCII+) and comparing their expression post-MI. **C,** Differential expression analysis of each cluster in the MI-D3 *Pcsk9^+/+^* group compared with the sham group (**Top**), and the MI-D3 *Pcsk9^−/−^* group compared with the MI-D3 *Pcsk9^+/+^* group (**Bottom**). **D,** Immunofluorescence (IF) staining of F4/80 (red) and Lyve1 (green) in AMI hearts. Scale bar, 20 µm. **E,** Quantification and statistical analysis of the percentages of Lyve1^+^ cells among F4/80^+^ macrophages (**Left**; *n* ≥ 4). Quantification and statistical analysis of the numbers of Lyve1^+^ macrophages (**Right**; *n* ≥ 4). **F,** IF staining of F4/80 (red) and CX_3_CR1 (green) in AMI hearts. Scale bar, 20 µm. **G,** Quantification and statistical analysis of the percentages of CX_3_CR1^+^ cells among F4/80^+^ macrophages (**Left**; *n* ≥ 4). Quantification and statistical analysis of the numbers of CX_3_CR1^+^ macrophages (**Right**; *n* ≥ 4). For **E** and **G,** each dot represents one mouse. All data are presented as the mean ± SEM. \**P* < 0.05.

### PDCMs drive cardiac regeneration

Macrophages play central roles in cardiac healing and tissue remodeling post-MI, influencing other cell types such as endothelial cells, smooth muscle cells, and fibroblasts to orchestrate the regenerative process.^8,26^ Our study explored the network of PDCMs with other cells in cardiac repair and regeneration. The data indicated that PDCMs exhibited advanced levels of cardiac muscle cell development, tissue development, and vasculogenesis compared with non-PDCMs based on scoring data (Figure 3A). During the regenerative phase of cardiac healing, myofibroblasts produce collagens, which help to maintain tissue structure and modulate the immunosuppressive functions of macrophages.^27,28^ Immunofluorescence and Masson’s trichrome staining demonstrated that PCSK9 deficiency was associated with enhanced myofibroblast density (α-smooth muscle actin) and collagen deposition in the injured heart, suggesting an improvement in wound healing capacity (Figure 3B and 3C). PCSK9 deficiency in myeloid cells increased microvascular density (CD31) and elevated *Pecam1* mRNA expression, indicating an enhanced angiogenic response in the ischemic region (Figure 3D and 3E). Thus, PCSK9-deficient cardiac macrophages promote advanced levels of coalition with other cardiac cell types, thereby improving wound healing in the infarcted heart. These results indicate that PCSK9 plays a pivotal role in modulating macrophage-mediated cardiac repair and regeneration.

**Figure 3.**
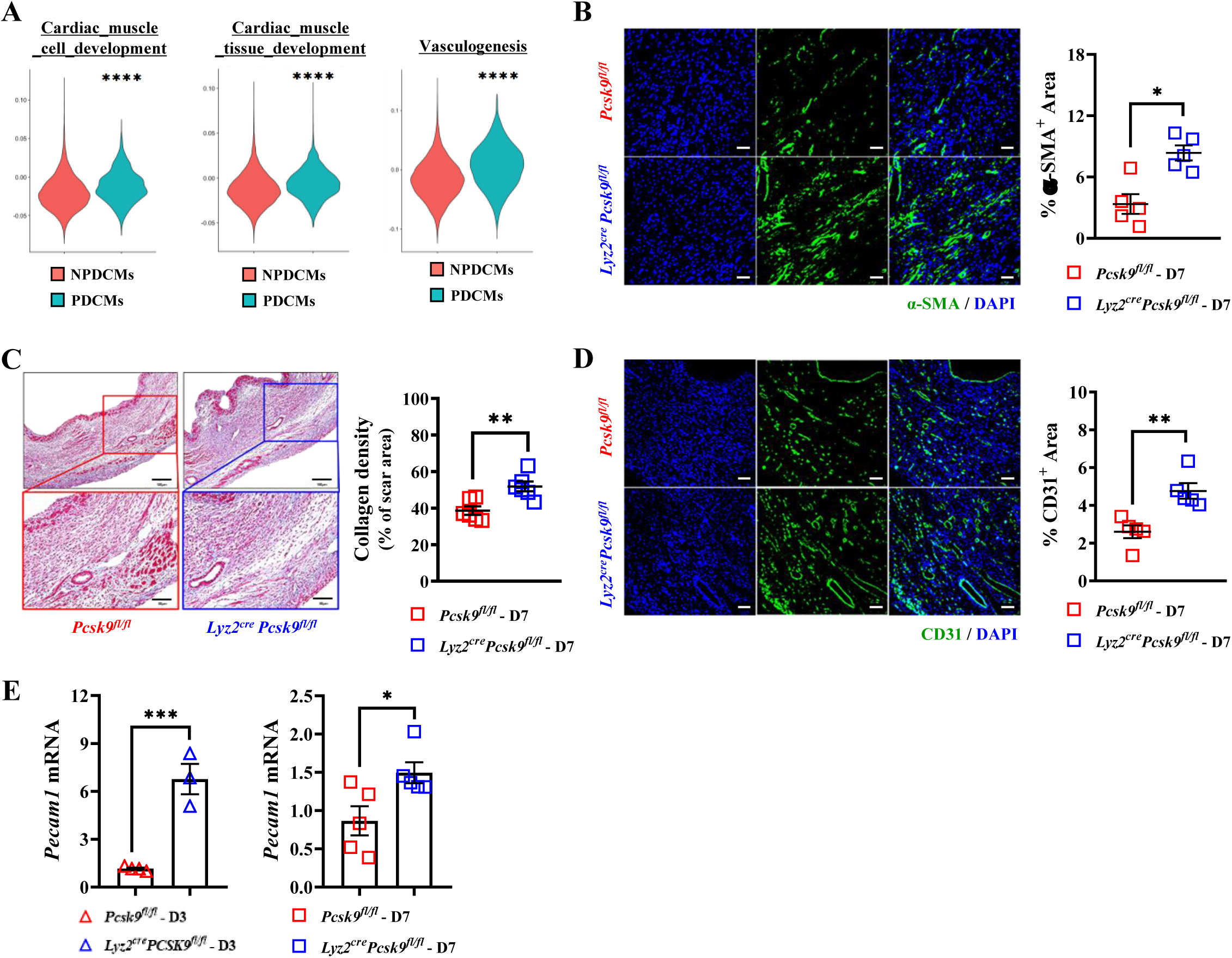
PDCMs enhance cardiac remodeling during the healing phase. **A,** Scoring data present enriched reparative roles such as cardiac muscle cell development, cardiac muscle tissue development, and vasculogenesis in PDCMs (Lyve1+, Cx3cr1+MHCII+) compared with the findings in NPDCMs (Res Mf, Ccr2+, Isg Mf) in the ischemic heart. **B,** Immunofluorescence (IF) staining of the myofibroblast density in infarcts from *Pcsk9^fl/fl^* and *Lyz2^cre^Pcsk9^fl/fl^* MI mice using α-smooth muscle actin (α-SMA)^+^ areas and quantitation graph at MI-D7. Scale bar, 20 µm (*n* = 5 per group). **C,** Masson’s trichrome staining was used to label collagen in infarcted hearts from *Pcsk9^fl/fl^* and *Lyz2^cre^Pcsk9^fl/fl^* MI mice and quantitation graph at MI-D7. Scale bar, 100 µm (**Top**), 50 µm (**Bottom**; *n* = 5 per group). **D,** Immunofluorescence staining of the infarct microvascular density in *Pcsk9^fl/fl^* and *Lyz2^cre^Pcsk9^fl/fl^* MI mice using CD31 and quantitation graph at MI-D7. Scale bar, 20 µm (*n* = 5 per group). **E,** qPCR analysis of the adhesion molecule *Pecam1* at MI-D3 (*n* ≥ 3) and MI-D7 (*n* = 5) in AMI hearts. Data in **B–D,** are representative of three independent experiments. For **B–E,** each dot represents one mouse. All data are presented as the mean ± SEM. ns, *P* > 0.05; \**P* < 0.05, \*\**P* < 0.01, \*\*\**P* < 0.001, \*\*\*\**P* < 0.0001.

### PDCMs exhibit enrichment in AP-1, promoting cardiac recovery in the infarcted area

PDCMs were identified as key regulators of the AMI repair process (Figure 2). To identify the critical regulators of PDCMs in the absence of PCSK9, Enrich R analysis was conducted. This analysis revealed elevated expression of AP-1 target genes in PDCMs following PCSK9 ablation (Figure 4A-4C). Immunoblotting of sorted CD45^+^Ly6G^−^CD11b^+^ myeloid cells and F4/80^+^ cardiac macrophages from the hearts of mice with AMI verified the upregulation of Lyve1, CX_3_CR1, and AP-1 (c-Jun) in *Pcsk9*-deleted mice compared with the findings in controls (Figure 4D, Figure VA and VB in the Data Supplement). Collectively, these observations suggest that PCSK9 deficiency promotes AP-1 activation in PDCMs, leading to enhanced healing signals and the inhibition of cardiac dysfunction.

**Figure 4.**
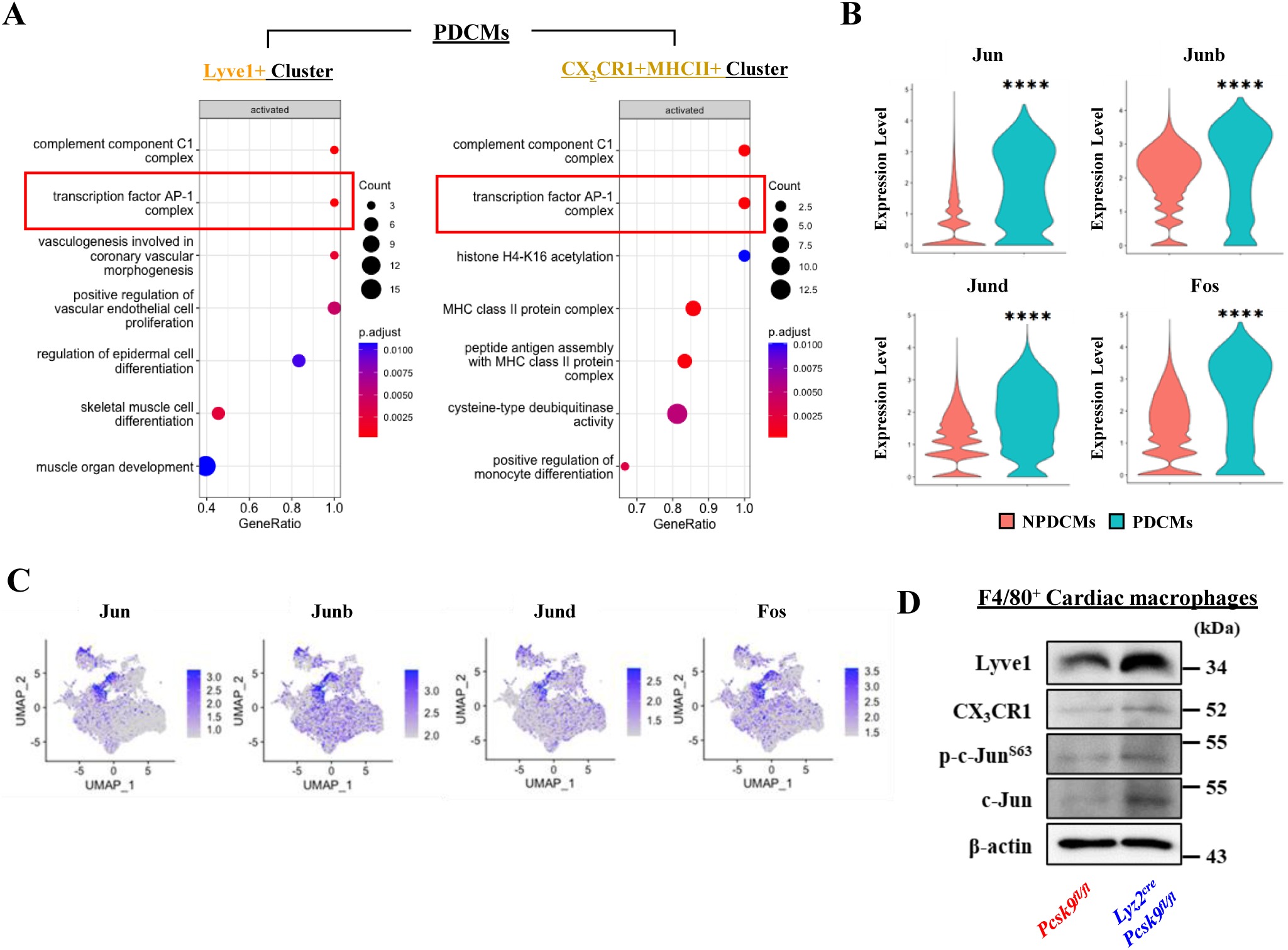
Enrichment of AP-1-related genes in the PDCMs. **A,** Genes upregulated in the absence of PCSK9 were assessed by Enrich R pathway analysis for PDCMs. **B,** Violin plots presenting the expression of representative markers of AP-1 expression for NPDCMs (Res Mf, Ccr2+, Isg Mf, Ly6C low, Ly6C int, Ly6C high, cDC1, cDC2, Unknown) and PDCMs (Lyve1+, Cx3cr1+MHCII+) in the ischemic heart. **C,** Feature plots depicting single-cell gene expression of AP-1 family genes. **D,** Immunoblot analysis of sorted F4/80^+^ cardiac macrophages for PDCMs from infarcted hearts on day 7. Data in **D,** are representative of three independent experiments. All data are presented as mean ± SEM. \*\*\*\**P* < 0.0001.

### AP-1/VEGF-C axis in PDCMs stimulates reparative Akt signaling in endothelial cells

AP-1 regulates macrophage activities, and it can promote the secretion of regenerative factors, such as VEGF.^29^ To ascertain whether PCSK9 deletion facilitates the secretion of regenerative components in the infarcted heart, we quantified VEGF family expression post-MI. The mRNA levels of *Vegf-a* and *Vegfr3* did not differ in the ischemia area, whereas *Vegf-c* and *Vegfr2* mRNA levels were elevated (Figure 5A, 5B and Figure VIA, VIB in the Data Supplement). Additionally, immunoblotting indicated that VEGF-C expression was upregulated in both sorted F4/80^+^ cardiac macrophages and CD11b^+^ cardiac myeloid cells from myeloid-specific PCSK9 deficient ischemic hearts on MI-D7 (Figure 5C and Figure VB in the Data Supplement). Although the assessment of improved VEGF-C activation and expression upon PCSK9 deficiency post-AMI suggest enhanced remodeling, these events could also reflect secondary effects related to the accentuation of restoration. To explore whether PCSK9 directly modulates VEGF-C synthesis by macrophages, we used BMDMs from *Pcsk9*-knockout mice. Secreted VEGF-C levels were higher in the BMDM(IL-4/IL-13)^ΔPCSK9^ supernatant under the AMI mimic conditions, as assessed by enzyme-linked immunosorbent assay (ELISA; Figure 5D).

**Figure 5.**
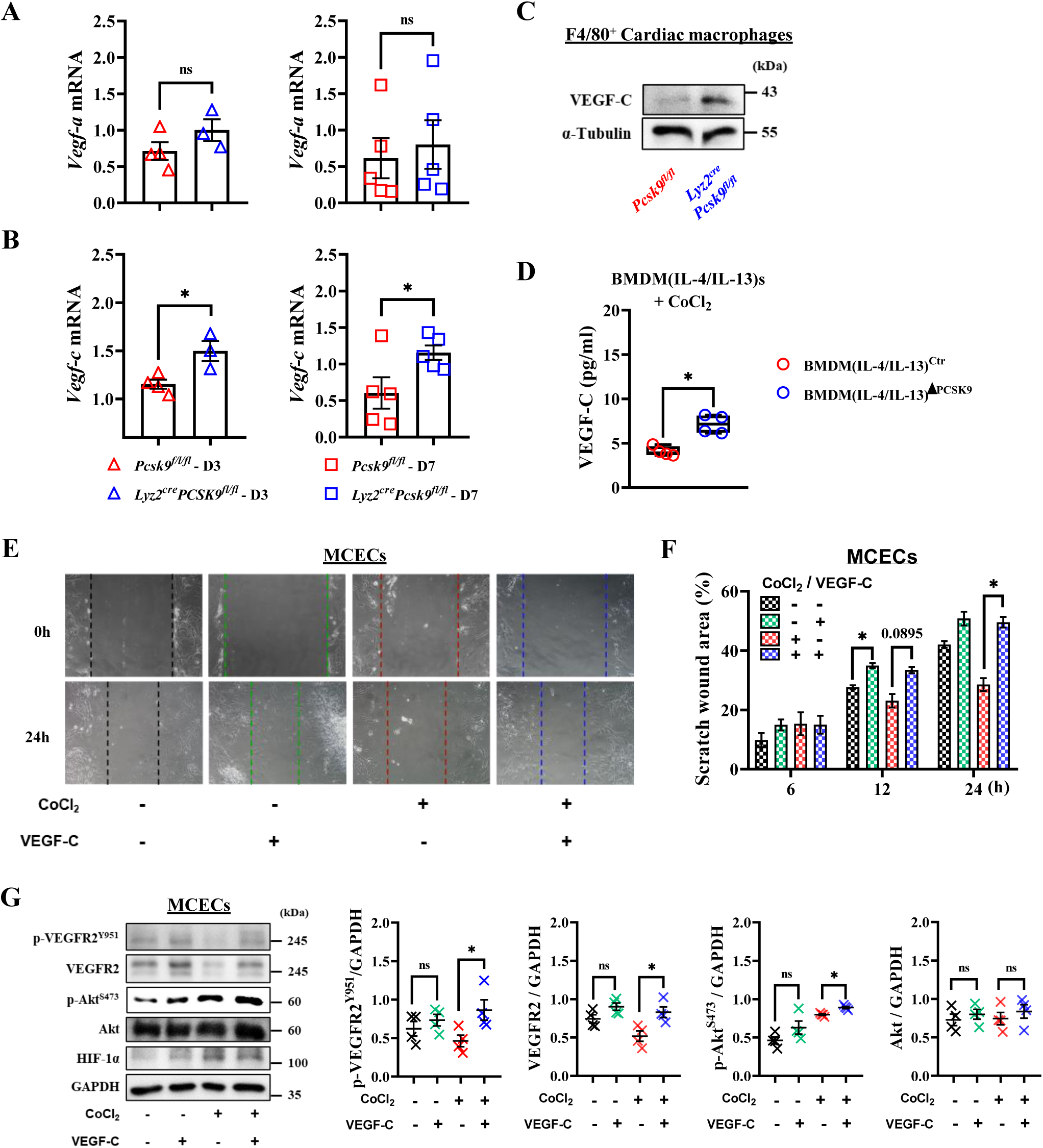
PDCMs improve cardiac repair through VEGF-C activation. **A** and **B,** qPCR analysis of vascular endothelial growth factor (VEGF) family members, *Vegf-a* **(A)**, *Vegf-c* **(B)** at MI-D3 (*n* ≥ 3) and MI-D7 (*n* = 5) in AMI hearts. **C,** Immunoblot analysis of sorted F4/80^+^ cardiac macrophages for VEGF-C from *Pcsk9^fl/fl^* and *Lyz2^cre^Pcsk9^fl/fl^* infarcted hearts on day 7. **D,** Secreted VEGF-C protein levels in supernatants from BMDM(IL-4/IL-13)s under ischemic conditions, assessed through ELISA (*n* = 4). **E-F,** Sorted and cultured Mouse Cardiac Endothelial Cells (MCECs) from mouse hearts. Representative images of the Scratch assay at 0h and 24h after treatment with CoCl_2_ and VEGF-C, respectively **(E)**. Quantification and statistical analysis of the scratch wound area at 6, 12, and 24h compared to 0h **(F)**. **G,** Immunoblot analysis for p-VEGFR2^Y951^, VEGFR2, p-Akt^S473^, and Akt expression and activation in MCECs after stimulation with CoCl_2_ and VEGF-C, respectively (*n* = 4). For **A,B,D,G,** each dot represents one mouse. Data in **E-G,** are representative of four independent experiments. All data are presented as mean ± SEM. ns; *P* > 0.05, \**P* < 0.05.

VEGF-C secreted from PDCMs as induced by AP-1 could bind to endothelial cell receptors, thereby stimulating Akt reparative signaling pathways to improve angiogenic activities.^30^ To determine whether VEGF-C stimulates Akt signaling pathways in endothelial cells during wound healing, we prepared and cultured mouse cardiac endothelial cells (MCECs) with VEGF-C and CoCl_2_. Using the scratch assay, we observed that VEGF-C enhanced MCEC migration under normal conditions after 12 h (black vs. green) and under ischemic conditions after 24 h (blue vs. red; Figure 5E and 5F). In human cardiac microvascular endothelial cells (HCMECs), VEGF-C also increased wound healing in the scratch assay (black vs. green at 24 h). Even under ischemic conditions, VEGF-C significantly improved proliferation after 12 and 24 h (red vs. blue; Figure VIIA and VIIB in the Data Supplement). Additionally, to verify the activation of the damage repair system, we analyzed VEGF-C–related receptor expression and Akt activation in MCECs. VEGF-C improved VEGFR2 activation and expression and increased Akt activation under hypoxic conditions (red vs. blue; Figure 5G). These findings suggest that the VEGFR2/Akt axis in cardiac endothelial cells is activated by VEGF-C secreted from PDCMs, promoting cardiac damage restoration.

### Identification of PDCMs features in myeloid cells from patients with CAD and human macrophages treated with a PCSK9 antibody

To investigate the relationship between immune cell heterogeneity and circulating PCSK9 levels in patients with vascular disease, we assigned patients into three groups: i) patients with hypertension not taking statins (non-CAD; Control), ii) patients using only statin therapy after CAD diagnosis, and iii) patients using statins and a PCSK9 antibody for at least 2 months after a CAD diagnosis. ELISA confirmed the increase in the plasma PCSK9 concentration in both the statin-only therapy and PCSK9 antibody treatment groups (Figure 6A). Flow cytometry revealed increases in myeloid cell and T cell counts among immune cell populations in patients treated with a PCSK9 antibody compared with the findings in patients treated with statins alone (Figure 6B, Figure VIIIA and VIIIB in the Data Supplement). The heterogeneity of myeloid cells, as observed *in vivo* and *in vitro*, was investigated in patients with CAD. Interestingly, we found that the myeloid cells from the PCSK9 antibody treatment group exhibited elevated expression of CX_3_CR1 and Lyve1 compared with the findings in the statin-only group, whereas CCR2 expression did not differ between the groups (Figure VIIIC in the Data Supplement). These results imply that PCSK9 antibody administration induces heterogeneity in myeloid cells in the blood. Furthermore, the plasma VEGF-C concentration was significantly higher in the PCSK9 antibody treatment group than in the control and statin-only groups, whereas VEGF-A levels did not differ among the groups (Figure 6C and Figure VIIID in the Data Supplement).

**Figure 6.**
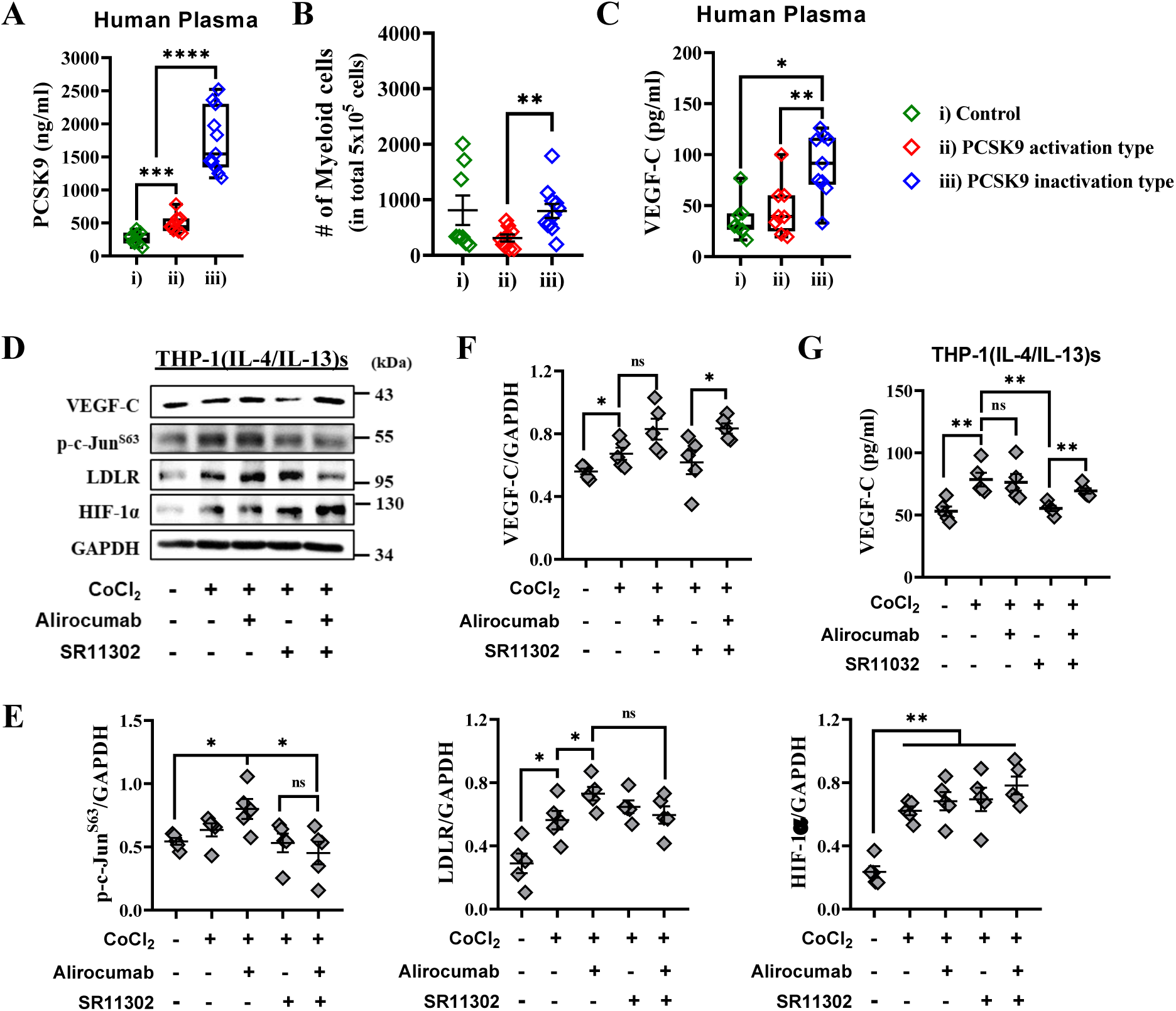
PCSK9 inhibitors alter myeloid heterogeneity in the plasma of patients with CAD and human macrophages. The study groups were as follows: **i)** non-CAD patients (hypertensive patients; Control group), **ii)** patients with CAD on statin therapy only, **iii)** patients with CAD on statin and PCSK9 antibody (Ab) therapy. PCSK9 Ab-treated patients with CAD were analyzed to explore changes in immune cell phenotypes. **A,** PCSK9 protein levels in plasma from each patient group as assessed by ELISA (*n* ≥ 5). **B,** Number of myeloid cells in each group analyzed by flow cytometry (*n* ≥ 5). **C,** VEGF-C protein levels in plasma from each patient group as assessed through ELISA (*n* ≥ 7). **D-F,** Immunoblot analysis of p-c-Jun^S63^, LDLR, HIF-1α, and VEGF-C expression in THP-1(IL-4/IL-13) cells pretreated with alirocumab (a PCSK9 inhibitor) and SR11302 (an AP-1 inhibitor) **(D)**. Quantitation of band density **(E-F)**. **G,** Secreted VEGF-C protein levels in supernatants from THP-1(IL-4/IL-13)s pretreated with alirocumab and SR11302 under ischemic conditions as assessed through ELISA. For **A-C,** each dot represents one person. Data in **D-F,** are representative of three independent experiments. All data are presented as mean ± SEM. ns; *P* > 0.05, \**P* < 0.05, \*\**P* < 0.01, \*\*\**P* < 0.001, \*\*\*\**P* < 0.0001.

Moreover, to demonstrate whether blocking AP-1 in human macrophages reduces VEGF-C levels, THP-1(IL-4/IL-13) cells were treated with CoCl_2_, alirocumab (a PCSK9 inhibitor), and SR11302 (an AP-1 inhibitor). As expected, HIF-1α expression increased under hypoxic conditions, LDLR expression improved following alirocumab administration^5^, and AP-1 (c-Jun) activation was blocked by SR11302 (Figure 6D and 6E). Notably, VEGF-C levels, which were reduced following AP-1 blockade alone, were restored by joint PCSK9 and AP-1 blockade (Figure 6D and 6F). ELISA indicated that although alirocumab treatment alone did not affect VEGF-C secretion, SR11302 reduced VEGF-C levels. Blocking both PCSK9 and AP-1 restored VEGF-C levels in the THP-1(IL-4/IL-13) cell supernatant (Figure 6G). Contrarily, the expression of VEGF-A, another regenerative factor, was not significantly affected by the inhibition of PCSK9 and AP-1 (Figure VIIIE in the Data Supplement). These findings indicate that AP-1 activation stimulates the secretion of VEGF-C, promoting cardiac protection and encouraging cardiac remodeling. The loss of PCSK9 in macrophages led to increased VEGF-C secretion, indicating that PCSK9 directly modulates VEGF-C synthesis by macrophages. Collectively, PCSK9-specific inhibition modulates myeloid cell phenotypes and elevates VEGF-C activation, which could represent a novel approach to enhancing cardiac remodeling and healing in MI and cardiovascular diseases.

## Discussion

The critical aspects of AMI pathophysiology are compounded by the intricate cellular mechanisms involved in the initiation and resolution of early-stage inflammation, as well as longer-term effects related to cardiac remodeling. Cardiac macrophages have emerged as key regulators during all stages after AMI, and distinct heterogeneity influences cardiac remodeling during the inflammatory and reparative phases. In this study, we observed that macrophages display PCSK9-dependent heterogeneity as a component of cardiac remodeling post-AMI, linking PCSK9 to reparative macrophage activity through an immunological mechanism independent of its role in cholesterol regulation. The function of PCSK9 in macrophage activation is (patho)biologically important, as genetic or antibody-dependent modulation of PCSK9 activity resulted in improved wound healing and cardiac remodeling, which were associated with a shift in macrophage heterogeneity toward a reparative phenotype. These findings highlight a novel function for PCSK9 in regulating the balance of macrophage heterogeneity and open therapeutic avenues toward expanding the clinical application of PCSK9 inhibitors.

Myeloid macrophages play critical roles in regulating inflammatory response and maintaining tissue architecture.^31^ We identified a novel subset of macrophages termed PDCMs, which facilitate healing and regeneration of infarcted tissue in mice lacking PCSK9 specifically in myeloid macrophages during the remodeling phase. Under hypoxic conditions, the phenotype of macrophages shifts from inflammatory to reparative earlier than observed under normoxic conditions, thereby initiating the repair process to promote myocardial recovery.^32^ Previous studies illustrated that myeloid cell subsets exhibit regenerative functions following mild traumatic brain injury^33^ and liver injury.^31^ Therefore, in the context of an infarcted heart, PCSK9-dependent reparative macrophages likely play a crucial role in successful heart repair. Targeting PCSK9 in cardiac macrophages could offer promising therapeutic approaches to mitigate tissue damage after AMI.

PCSK9 regulates plasma LDL-c levels by promoting the lysosomal degradation of LDLR. Notably, our findings revealed that PCSK9 deficiency in cardiac myeloid cells attenuates AMI independently of LDL-c levels. The regulatory mechanism of PCSK9, beyond its role in cholesterol homeostasis, and its association with human diseases remain elusive. In hepatocytes, but not cardiac cells, cyclase-associated protein 1 (CAP1) has been identified as a key factor that binds to PCSK9 for caveolae-dependent endocytosis.^4^ Inhibition of PCSK9–CAP1 binding has been found to suppress atherosclerotic inflammation independently of LDLR.^34^ Other research demonstrated that PCSK9 targets LDLR-related protein 5 (LRP5), which mediates lipid uptake in macrophages under atherosclerosis.^16^ These findings suggest that proteins other than LDLR are also involved in PCSK9-related mechanisms in coronary arteries.

AP-1, well known for its role in wound healing in skin fibroblasts^35^, does not have a well-defined role in cardiac macrophages concerning ventricular remodeling post-MI. Cardiovascular pathophysiology increases cardiac workload as a compensatory mechanism, potentially increasing the risk of heart failure.^36^ Under stress, the AP-1 component JunD has been demonstrated to reduce hypertrophy.^37,38^ Conversely, *in vivo* studies revealed that hearts lacking Jun develop fibrosis and exhibit increased myocyte apoptosis, potentially leading to dilated cardiomyopathy.^39^ These findings suggest that Jun is essential for initiating a transcriptional program that adapts to increased workload in the heart. Our study also observed increased expression of Jun family members in PDCMs after AMI, suggesting potential roles in cardiac adaptation and remodeling following injury. Further experiments are necessary to clarify the factors that stimulate AP-1 during cardiac remodeling upon PCSK9 deficiency.

Reducing LDL levels can mitigate cardiovascular risk, with statins, ezetimibe, and PCSK9 inhibitors playing pivotal roles in upregulating LDLR.^40^ However, these drugs can paradoxically elevate plasma PCSK9 levels^41,42^, potentially leading to secondary risks. Additionally, reports noted that statin treatment alone can significantly reduce plasma VEGF concentrations^43^ and suppress vasculogenesis.^44^ In this study, both plasma VEGF-A and VEGF-C levels were unchanged in the plasma of patients with CAD using statins alone. Therefore, a second agent, such as ezetimibe or a PCSK9 inhibitor, is advised for patients at high cardiovascular risk or those intolerant to statins. Recent studies demonstrated that combining statins with PCSK9 inhibitors is more effective than combining statins with ezetimibe in reducing the risks of MI and stroke.^45^ Our experiments suggested that adding PCSK9 antibody to statins in patients with CAD increases the number of reparative myeloid cells with PDCM-like properties and raises plasma VEGF-C levels, indicating a potentially favorable prognosis.

In conclusion, targeting myeloid-PCSK9 could led to favorable therapeutic outcomes because of its cell type- and tissue-specific activities. Indeed, myeloid-PCSK9 deficiency in mice post-AMI and PCSK9 antibody-treated patients with CAD exhibited features of PDCMs promoting enhanced cardiac remodeling and disease alleviation. Our findings demonstrate, for the first time, a potential clinical strategy involving myeloid-specific PCSK9 for protecting against MI and other vascular diseases beyond lowering LDL-c.

## Data Availability

scRNA-seq data for this project have been deposited at NCBI?s Gene Expression Omnibus (GEO) and GSE number is pending.

## Acknowledgements

S.H.M., H.W.K., S.H.P., and G.T.O. designed the experiments. S.H.M. planned and performed most of the experiments, and help from N.H.Y., K.I.C., H.J., and Y.S.N. also contributed to several experiments. H.W.K. and S.H.P. conducted the scRNA-seq data analysis and validation. J.J., S.J., S-K.S., S.S., J.S., H.Y.K., N.H.Y., and H.J. advised and commented on the interpretation of the work. N.G.S. developed and provided *Pcsk9*^−*/*−^*, Pcsk9^fl/fl^*mice and discussed of the work. W.K.Y helped measure blood lipid levels. S-J.L. and C.J.L. contributed to the selected and recruited patient study participants. S.H.M. and G.T.O. wrote the manuscript (original draft), and S.H.M., H.W.K., S.H.P., and G.T.O. reviewed and edited the manuscript. All authors read and approved the manuscript.

The apparatus including LSM780 and LSM880 NLO (Zeiss), LSRFortessa (Becton Dickinson), and FACSAria Fusion (Becton Dickinson) at Ewha Fluorescence Core Imaging Center were utilized for major experiments.

## Nonstandard Abbreviation and Acronyms

AMI: acute myocardial infarction
CAD: coronary artery disease
PCSK9: pro-protein convertase subtilisin/kexin type 9
LDL: low-density lipoprotein
PDCM: PCSK9-dependent cardiac macrophages
MCECs: mouse cardiac endothelial cells
HCMEC: human cardiac microvascular endothelial cells

